# Comparison of Pandemic Intervention Policies in Several Building Types Using Heterogeneous Population Model

**DOI:** 10.1101/2021.07.15.21260564

**Authors:** Teddy Lazebnik, Ariel Alexi

## Abstract

In a world where pandemics are a matter of time and increasing urbanization of the world’s population, governments should be prepared with pandemic intervention policies (IPs) to minimize the crisis direct and indirect adverse effects while keeping normal life as much as possible. Successful pandemic IPs have to take into consideration the heterogeneous behavior of individuals in different types of buildings and social contexts. In this study, we propose a spatio-temporal, heterogeneous population model and *in silico* simulation to evaluate pandemic IPs in four types of buildings - home, office, school, and mall. We show that indeed each building type has a unique pandemic spread and therefore a different optimal IP. Moreover, we show that temporal-based IPs (such as mask wearing) have a similar influence on the pandemic spread in all four building types while spatial-based IPs (such as social distance) highly differ.

## 1 Introduction

Humanity has experienced multiple epidemics over the centuries which caused significant mortality, economic losses, and political shifting [10]. In the last fifty years, the world has experienced four large-scale outbreaks of pandemics: HIV/AIDS, Seventh Cholera, 2009-flu, and Coronavirus [8]. The question of how policymakers can control a pandemic spread is becoming increasingly more relevant as urbanization in the developing world is bringing more people into denser neighborhoods, which increases the speed at which new infections are spread [28]. Moreover, globalization has facilitated pathogen spread among countries through the growth of trade and travel [51]. These socioeconomic processes keep the infectious disease outbreaks nearly constant, even with modern medicine, technology, and government awareness [8].

Thus, the preparedness of policymakers is a necessary step to ensure the ability of a country in handling the next pandemic. A useful tool to obtain data-driven decisions such as lockdowns [1], artificial job separation [27], school-work duration [26], mask wearing [34] and others [12] pandemic intervention policies (PIPs) are epidemiological-mathematical models which allows to investigate the influence of these PIPs in multiple scenarios. A large portion of the epidemiological models are based on the SIR model [22] with different extensions related to biological, economical and spatial properties of the disease that one is aiming to model [11, 17, 52].

Nevertheless, these models treat individuals homogeneously, assuming everybody have the same biological properties, located in the same places during the day, and meet each other randomly or by a static network of connections [26, 49]. To address this shortcoming, we developed a spatio-temporal social-epidemiological model which treat each individual heterogeneously by allowing a unique walk between the locations (rooms) based the individual’s social role and “personality”. The proposed model describes the way populations move around a building using a graph-based spatial model and the pandemic spread using an extended SIR model. In addition, we developed a computer simulation that provides an *in silico* tool for evaluating the performance of four PIPs over four different types of buildings: home, office, school, and mall. The proposed model allows a more accurate investigation of the epidemiological dynamics in relatively small population sizes. Using the model, we show that temporal-based PIPs behave identically if the population is dense enough while spatial-based PIPs behave differently in these building types. Moreover, we show that the PIPs’ dynamics in the *home* building type is chaotic due to low density of population except of the social distancing PIP which significantly reduce the pandemic spread.

This paper is organized as follows: In Section 2, we provide background about mathematical models for pandemic spread followed by graph-based population interaction models and a review of PIPs. In Section 3, we introduce our spatio-temporal social-epidemiological model. In addition, we provide an agent-based approach [30] to simulate the model’s dynamics. In Section 4, we present four PIPs used to control the pandemic spread. In Section 5, we present the implementation of the model for the COVID-19 pandemic and data of buildings from Israel, followed by sensitivity analysis of the PIPs for the different building types. In Section 6, we discuss the main advantages and limitations of the model. In Section 7, we briefly describe the possible usages of the model for policymakers in future pandemics.

## 2 Background

Historical records from the last few hundred years show that pandemic caused significant political shifting, economical losses, and mortality [10]. In particular, in the 20th century (1918) the influenza pandemic struck the world and killed an estimated 50 to 100 million individuals worldwide [10] and in 2020 the coronavirus (COVID-19) pandemic was declared by the World Health Organization (WHO) as a public health emergency of international concern and killed millions [44].

One approach to allow policymakers to better manage pandemics is using mathematical models and computer simulations. Indeed, multiple models are shown to be efficient in obtaining data-driven decisions such as artificial job separation [27], lockdowns [1], masks wearing [34], school-work duration [26], and others PIPs [12]. These mathematical models can be divided into two main groups. First, models aim to predict the different parameters such as the total death and peak in hospitalized individuals, given historical data [45, 36]. Second, models that aim to analyze and optimize PIPs [53, 13].

Models related to the second group are usually focusing on large populations such as cities [13], countries [45, 18], and even the entire world population [36]. As such, they ignore the small, daily interactions of the individuals in the population that influence the overall pandemic spread dynamics [15]. The world’s population is growing [42] and is concentrated in cities in general and megalopolis cities in particular [24]. As a result, most of the individuals spend most of their time indoors (22-23 hours a day on average) [14].

We present an overview of epidemiological models, in which, general and spatio-temporal epidemiological models are shown as a method for the heterogeneous indoor epidemiological model. Afterward, several graph-based population interactions models are presented, the describe studies where using this representation allowed us to investigate the merged behavior of a population from the individual level. Finally, several studies outline possible PIPs.

### 2.1 Epidemiological models

Mathematical models of epidemiological dynamics usually represent the transformations of individuals in the population between several epidemiological states [52, 11, 26, 5]. These models can be roughly divided into two main groups: diseases with short-term and long-term immunity memory when more often than not an immunity memory is considered long-term if it is longer than the average duration of two infection waves in the population. For example, the influenza is short-term immunity memory disease since it occurs on average once a year when the immunity memory is shorter [20].

The group of short-term immunity memory diseases can be represented by the Susceptible-Infected-Susceptible (SIS) model, where susceptible (S) individuals are infected on average at a rate *β* relative to the size of the infected individuals sub-population, and infected individuals (I) recover at a rate *γ* and become susceptible (S) again [2]. The SIS model is represented by the following non-linear ODEs:

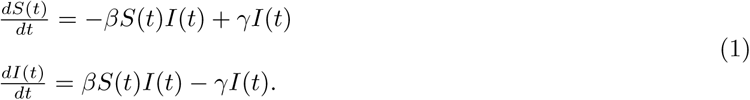

In the context of indoor dynamics with a relatively short time frame, individuals develop and retain the immune system needed to be safe from re-infection [32]. Therefore, the epidemiological dynamics are better fit to the long-term immunity memory diseases regardless of the type of the disease itself. Therefore, the model takes the form of the Susceptible-Infected-Recovered (SIR) model [22], represented as a system of ODEs where susceptible (S) individuals are infected on average at a rate *β* relative to the size of the infected individuals sub-population and infected individuals (I) recover at a rate *γ* and become recovered individuals that cannot be infected again. The SIR model is represented by the following non-linear ODEs:

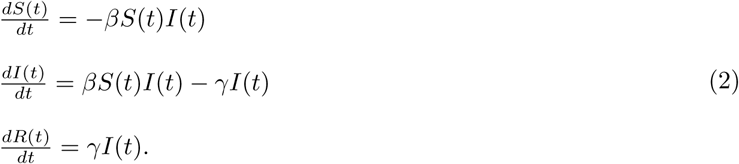

While the SIR model is widely used due to its simplicity, which is commonly used to obtain an initial estimation of the pandemic behavior, its simplicity is also its shortcoming as it fails to represent the complexity of the pandemic spread dynamics [36]. Therefore, several extensions of the SIR model are proposed to better represent the spread dynamics:

First, the mortality due to the pandemic defines a state *D*. This state plays a meaningful role in the dynamics as it removes individuals from the interaction, besides allowing to investigate and predict the portion of individuals that die due to the pandemic [53].

Second, adding an exposure state (E) which represents the period between the time a susceptible individual is infected and until it starts to be infectious to others [45]. The exposed state better represents the biological settings as many diseases go through a phase of incubation [25, 47].

Third, individuals experience diseases in a range of severity [45]. Therefore, by dividing the infection group, *I*, into two degrees of infection severity: symptomatic, *I*^*s*^, and asymptomatic, *I*^*a*^, it is possible to obtain a more accurate representation of the social and epidemiological dynamics as well as to fine-tune the infection rates as different severity degrees have corresponding infection rate.

Fourth, data from several epidemiological studies show that children and adults transmit diseases at different rates and have different recovery duration [19, 43]. As a result, it is possible to divide the population into two age groups, adults, and children, such that each of the values of the transformation between epidemiological states within and between groups are different [9].

Fifth, the places where individuals spend their time during the day affect the pandemic dynamics by changing the rate of infection. Indeed, in the case of COVID-19, Viguerie et al. [46] showed that Spatio-temporal SIR-based models better predicted the COVID-19 spread in the Italian region of Lombardy. Their version of the spatial dynamics assumes the static distribution of the population over the course of the day and does not take into consideration the unique dynamics of a different location as is possible by using a graph-based spatial model [26].

### 2.2 Graph-based population interactions models

Individuals movement around in buildings, occupancy and interactions with devices are influenced by variables in three main categories: environmentally-related, time-related, and random. The environmentally-related variables include a physical aspect related to the building’s characteristics and location. Solar orientation, envelope, building layout, and local climate are some examples of environmentally related factors. The time-related variables comprehend the occupants’ routine. In that manner, occupancy and interactions with devices and individuals in buildings are influenced by time of day and day of week [4].

Noakes et al. [38] modified the classic SIR model, taking into consideration several physical properties of airborne diseases in a close space based on the physical airborne infection model proposed by Riley et al. [41]. The model is able to evaluate the effects of room size, occupancy and ventilation conditions on the number of new infections which makes it more accurate than the general-proposed SIR model. Nevertheless, the model assumes the population is located in the room during the entire dynamics which is a poor approximation of the real movement dynamics of individuals over relatively long periods (days, weeks, and months) in which usually pandemic prediction takes place. Therefore, we assume individuals move between rooms over time, which changes the infection rate in rooms accordingly.

Several more studies investigated the epidemiological dynamics of people as a network of interaction represented using a graph-based model. Hau et al. [17] proposed an SEIR-based model (E-exposed) for sexually transmitted diseases where the interactions between individuals happened randomly on a bipartite (male and female nodes) static graph. Similarly, Wang et al. [49] proposed an SIS model where each individual is represented as a node in a static graph that has between 1 and *k << N* edges randomly set (where *N* is the size of the population). The assumption of a static graph over time introduces an error to the simulation as unplanned interactions (for example, using an elevator with unfamiliar people), changes in contacts (for instance, when replacing a workplace or when meeting new members in ones sports team), and other factors make edges of the graph dynamic. In the proposed model, we consider these dynamics using the walk between rooms of a building which in its turn define the interactions between individuals at any point in time.

### 2.3 Pandemic intervention policies

Governments across the world are facing the task of selecting suitable intervention strategies to cope with the effects of pandemics. This is a highly challenging task, since harsh measures may result in an economic collapse while a relaxed strategy might lead to a high death toll. Pandemic intervention policies (PIPs) are aimed to optimize the trade-off between the number of infections (and therefore deaths) and the socioeconomic costs.

Non-pharmaceutical intervention (NPI) policies are actions, apart from getting vaccinated and taking medicine, that individuals and communities can take to slow the spread of a pandemic. There are multiple NPI policies such as school-work duration [26], lockdown [27], mask wearing [52] and others. Zhao et al. [53] proposed an extension to the SEIR model where the susceptible population is separated into two groups: individuals not taking infection-prevention actions and individuals taking infection-prevention actions as an NPI policy. The authors introduce a stochastic element to the SEIR model making it more robust for social changes that happened during the epidemic. Di Domenico et al. [13] used data from March 17 to May 11 (2020) in Île-de-France with a stochastic age-structured transmission extension of the SEIR model integrating data on the age profile and social contacts of four age-based classes. The authors investigated the influence of average social distancing on the duration of the pandemic and the total number of infected individuals.

On the other hand, pharmaceutical intervention policies are clinical actions such as vaccination and taking medication. Moore et al. [35] investigated the influence of age-based vaccination on the changing levels of infection based on epidemiological data from the UK using the SIR model for the case of COVID-19.

## 3 Model definition

The model can be mathematically described using an interaction between two sub-models: epidemiological (temporal) ODE-based and social (spatial) graph-based sub-models. It is difficult to numerically solve this representation due to the noncontinuous accrual as a result of the spatial dynamics (population mobility in the building). In addition, it is also difficult to use this representation to obtain analytical results, due to the nontrivial integration of ODE and graph theories and the large-scale representation of the dynamics it yields. Therefore, we proposed an agent-based approach to simulate the proposed model. The examined PIPs are treated as an additional layer to the model by modifying several attributes of the model.

### 3.1 Epidemiological (Temporal) sub-model

The model considers a constant population with a fixed number of individuals *N*. In the context of an indoor pandemic, the time horizon of interest is relatively short and therefore the native population growth can be neglected. Each individual belongs to one of seven groups: susceptible (*S*), asymptomatic exposed (*E*^*a*^), symptomatic infected (*E*^*s*^), asymptomatic infected (*I*^*a*^), symptomatic infected (*I*^*s*^), recovered (*R*), and dead (*D*) such that *N* = *S* + *E*^*a*^ + *E*^*s*^ + *I*^*a*^ + *I*^*s*^ + *R* + *D*. Individuals in the first group have no immunity and are susceptible to infection. When an individual in the susceptible group (*S*) is exposed to the pathogen, the individual is transferred to either the asymptomatic exposed group (*E*^*a*^) or symptomatic exposed group (*E*^*s*^) at a rate corresponding to the average interaction between infected individuals and susceptible individuals. The individuals stay in the symptomatic, asymptomatic exposed group on average *ξ*^*s*^, *ξ*^*a*^ days, respectively, after which the individual is transferred to the symptomatic, asymptomatic infected group. Afterward, the individual stays in the symptomatic infected group on average *γ*^*s*^ days, after which the individual is transferred to the recovered group (*R*) or the dead group (*D*). Therefore, a rate of (1 − *ψ*) of symptomatic infected individuals remain seriously ill or die while others recover. All asymptomatic infected individuals stay in the infected *I*^*a*^ group on average *γ*^*a*^ days, after which the individual is transferred to the recovered group (*R*). The recovered individuals are again healthy, no longer contagious, and immune from future infection.

The population is further divided into two age classes: adults and children, because these groups experience the disease in varying degrees of severity, have different infection rates, and different social roles. Individuals below age *A* are associated with the *children* age-class while individuals in the complementary group are associated with the *adult* age-class.

By expanding the designation to two age-classes, we let 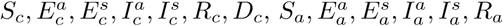, and *D*_*a*_ represent susceptible, asymptomatic exposed, symptomatic exposed, asymptomatic infected, symptomatic infected, recovered, and death groups for children and adults, respectively such that

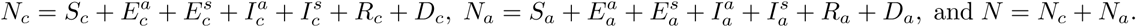

The epidemiological dynamics (e.i., the SEEIIRD model) are described in detail in Eqs. (S1-S14) in the supplementary material. A summary of Eqs. (S1-S14) is shown in Eq. (3).

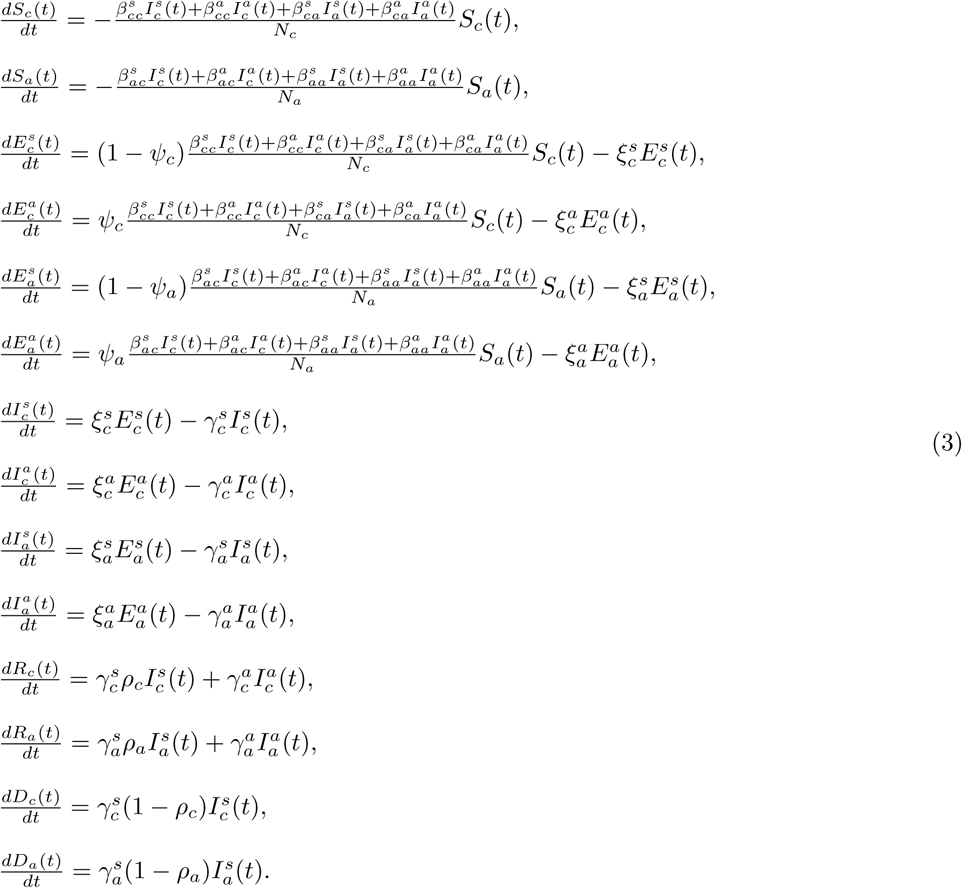

However, we treat the coefficients as probabilities rather than rates as they represent the probabilities for state transfer in the individual level [11]. This results in making Eq. (3) a non-linear, first order, fully dependent, stochastic ODE system with 14 states. A schematic view of the transformation between the epidemiological model’s state is shown in Fig. 1.

**Figure 1:**
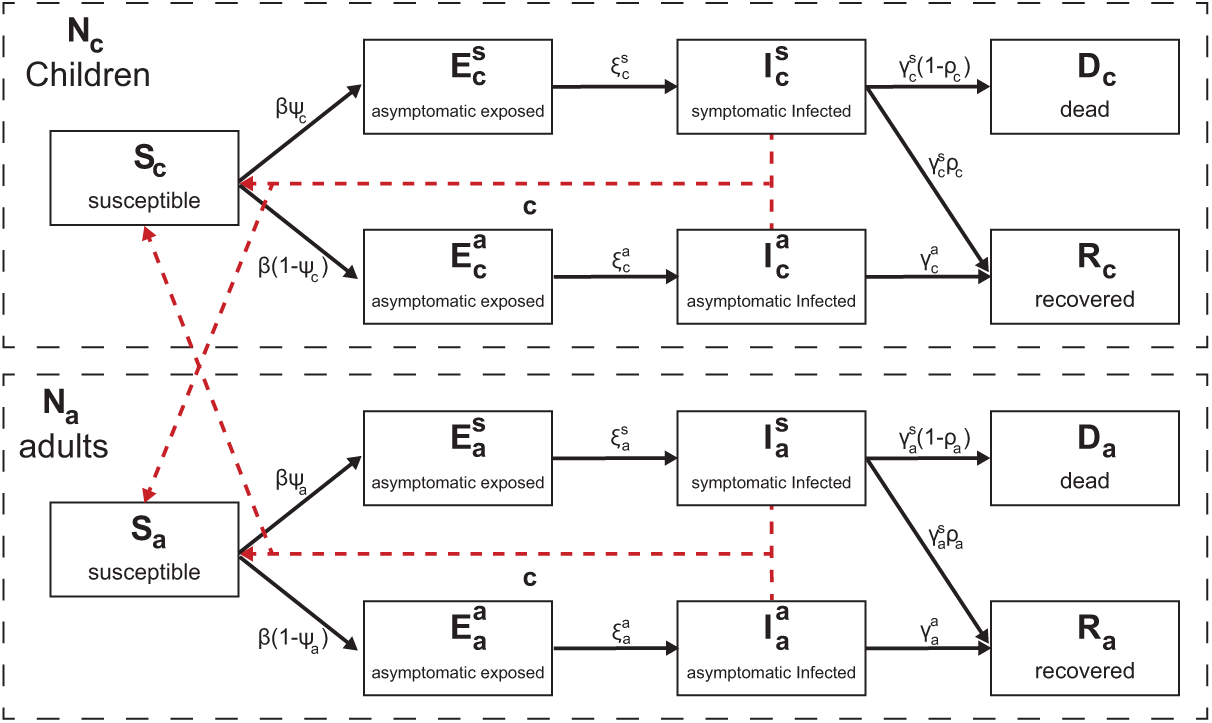
A schematic view of an individual’s transformation between epidemiological states. The solid arrows indicate a state transformation while the dashed arrows indicate infection interactions.

### 3.2 Social (Spatial) sub-model

The spatial sub-model is a graph-based model *G* = (*V, E*). The population *N* from the temporal dynamics is allocated in some distribution to the nodes of an undirected, connected graph (*G*). The graph is defined according to the rooms *V* (nodes) of a building and the connections *E* (edges) between them which are defined if there is a door between two rooms. For each step in time, the population on the graph is moving to one of the nodes in the graph or staying in the same node, according to the inner moving policy each agent has in addition to a global policy which overtakes the decision in the case of a conflict between the two policies. The transition between any two nodes is assumed to be immediate and that everybody is following the same clock. Between each population movement on the graph, the temporal sub-model is performed simultaneously on all the nodes of the graph.

Therefore, based on the *Wells–Riley* equation [41] and the extension proposed by Noakes et al. [38], in each room the infection probability is computed using the formula:

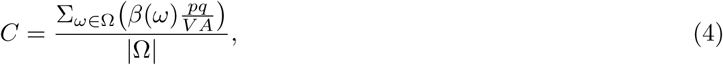

where *C* is the probability a susceptible individual will be infected by spending time in the same room as infected individuals, Ω is the set of all sub-groups of susceptible *S* and infected (*P*) sub-populations. For example,

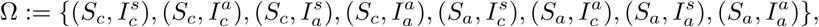

for the proposed epidemiological model. *β*(*ω*) is the infection probability of the pair of sub-populations *ω* in general, *p* is the average pulmonary ventilation rate of the susceptibles (*m*^3^*/t*), *q* is the quanta production rate per infector (*t*^−1^), *A* is the ventilation rate in air changes per hour (*t*^−1^), and *V* is the room volume (*m*^3^).

Eq. (4) holds true in the case of airborne disease. In the case the disease is not airborne (like sexual diseases), the probability *C* equals the coefficient of the *SI* term in the epidemiological (temporal) model (see Section 3.1).

### 3.3 Numerical simulation

Due to the non-linear and large-scale (14V equations) nature of the system in addition to the noncontinuous behavior as a result of the population mobility between the rooms - it is hard to numerically solve the system in both a stable and fast way as current ODE solvers struggle in such a case even on small-scale systems [31, 3].

Therefore, we take advantage of the P-system model [40] to simulate epidemiological and social dynamics as an agent-based simulation is a powerful tool to simulate complex social systems [30]. Specifically, we extend the model proposed by [6]. First, let us define the system as an instance of a P-system model. A P-system model *PM* is defined as a tuple of three elements *P*_*s*_ := (*P, M, I*) where *P* is a set of finite state machine agents, *M* is a set of locations (originally, membranes) over which the population is distributed, and *I* : (*p*_*i*_, *p*_*j*_) → (*p*_*i*_, *p*_*j*_) is a pair-wised interaction protocol as a function between two agents *p*_*i*_, *p*_*j*_ ∈ *P* such that *i* ≠ *j* changes their states. The location *m* ∈ *M* that the agent is located at a given time is part of the state definition of the agent. At each point in time, the population is randomly divided into pairs such that agents in a pair are constrained to be located in the same location *m* ∈ *M*. Afterward, the interaction protocol *I* is performed on each pair.

We introduce an extended P-system definition. An EP-system is defined as a tuple *EP*_*s*_ := (*P, G, I*_*p*_, *S*_*p*_, *M*_*p*_) where *P* is a set of timed finite state machine agents where the inner clock measure the time pass from the last state change, *G* = (*M, E*) is an underacted, connected graph over which the population is distributed, *I*_*p*_ : (*p*_*i*_, *p*_*j*_) → (*p*_*i*_, *p*_*j*_) is a pair-wised interaction protocol as a function between two agents *p*_*i*_, *p*_*j*_ ∈ *P* such that *i* /= *j* changes their states, *S*_*p*_(*p*) → *p* is a mono-wised spontaneous protocol as a function that changes the state of an agent based on the inner clock and current state, and *M*_*p*_(*G, P*) → *P* is a movement protocol as a function from the graph *G* and the population *P* and return a new distribution of the population *P* in the graph *G*. At each point in time, the population is randomly divided into pairs such that agents in a pair are constrained to be located in the same location *m* ∈ *M*. Afterward, the interaction protocol *I* is performed on each pair. Then, the spontaneous protocol *S*_*p*_ and the movement protocol *M*_*p*_ are performed on each agent in the population.

Therefore, let *M* be an EP-system which represents the model defined in Sections 3.1, 3.2. Namely, *P* is a set of individuals, *G* is the graph of rooms in the building, connected by edges, which is a physical way to go from one room to another (usually, via a door). The interaction protocol *I*_*p*_ implements the infection dynamics of the SEEIIRD model stochastically:

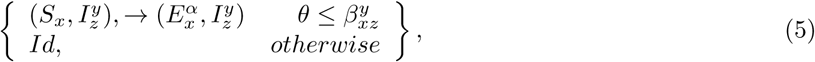

where *θ* ∼ *U* [0, 1] is the chance that the current infection succeeds, 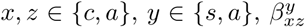 is the average probability that a susceptible *x* age group individual would be infected from a *z* age group individual with *y* infection severity, and *α* ∈ {*s, a*} is set to *s* at a probability *ϕ*_*x*_ and *a* otherwise. The spontaneous protocol *S*_*p*_ implements the infection and recovery dynamics using the inner clock (T):

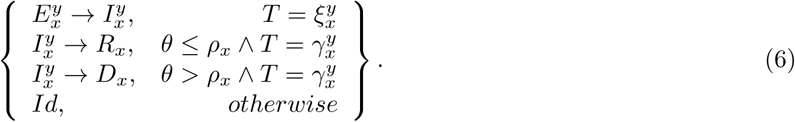

Finally, the movement protocol *M*_*p*_ is a general purpose function which is defined as part of a specific policy.

## 4 Pandemic Intervention Policies

Pandemics have negatively impacted many aspects of humanity’s existence, causing massive unrest around the world with significant loss of life. Policymakers are forced to aim to execute PIPs in the form of non-pharmaceutical intervention (NPI) policies such as social distancing and masks and pharmaceutical intervention (PI) policies such as vaccination to control the epidemic. These PIPs can be divided into two main groups: temporal and spatial. The temporal PIPs modify the individual or population’s properties related to the transformations between the epidemiological stages (see Section 3.1) while spatial PIPs modify the individual or population’s properties related to the walks of the population in the topology.

### 4.1 Spatial-based policies

In the model, a spatial-based policy fully or partially overrides the walk on the topology dynamics each individual has. Specifically, in each step in time, the policy modifies the walk dynamics of all individuals in the population at once followed by regular heterogeneous walk behavior of each individual independently, according to these modifications.

#### 4.1.1 Isolatation of symptomatic infected individuals

Symptomatic (exposed and infected) individuals produce clinical signs which are relatively easy to measure (for example, increased body temperature), and it is recommended to isolate these individuals from the rest of the population to reduce the infection rate. Since the measuring process requires effort in the form of manpower and technical means (for instance, a distance thermometer), it is limited by the availability of these factors.

Therefore, the isolation of symptomatic infected individuals (ISII) policy is realized as follows. Every *τ* ∈ [0, ∞) steps in time, all individuals at this point in a portion *σ* ∈ [0, 1] of the rooms of the building are tested. Individuals that belong to the symptomatic exposed (*E*^*s*^) symptomatic infected (*I*^*s*^) are isolated out of the building. In addition, at each check, individuals that were isolated in a previous check are re-tested and if they are recovered (*R*) they are allowed to return to the building and continue their original behavior.

#### 4.1.2 social distancing

During an airborne type pandemic, the instruction for individuals is to keep social distancing to reduce the infection rate. Social distancing reduces the infection rate as the sum of possible infections in each room ∑_*ω*∈Ω_*β*(*ω*) is decreasing (see Eq. (4)).

Therefore, the social distancing (SD) policy was realized as follows. There is a probability *χ* ∈ [0, 1] that an individual overrides its original walk dynamics with a walk to a neighboring room that optimizes the SD, where SD in a room *i* is defined as follows:

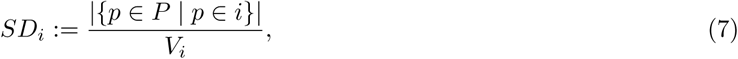

where *V*_*i*_ is *i*’s room volume.

### 4.2 Temporal-based policies

In the model, a temporal-based policy modifies the spontaneous and interaction protocols of the individuals of the population, and does not change their behavior.

#### 4.2.1 Mask wearing

During an airborne type pandemic, wearing masks reduces the rate of infection in the event of an encounter between individuals [29]. However, masks have several levels of protection that differ according to their materials [39].

Therefore, the mask wearing (MW) policy was realized as follows. When two individuals such that one of them is infected and the other is suspicious interact, there is a probability *C* (see Eq. (4)) that the suspicious individual would be infected. If one of the sides wears a mask with quality *α* ∈ [0, 1], the infection probability becomes *αC*. If both sides wear a mask with quality *α* ∈ [0, 1], the infection probability becomes *α*^2^*C*. The portion of individuals that are wearing masks all the time is marked by Γ ∈ [0, 1].

#### 4.2.2 Vaccination

Vaccination is known to be the golden pandemic intervention policy [23]. Vaccination of the entire population for large populations is a complex, time and resource-consuming task [23]. As a result, it is common that only a portion of the population is vaccinated. In addition, a vaccine is not a silver bullet since it is effective only for a portion of the time and vaccinated individuals may be infected anyway.

Therefore, the vaccination (V) policy was realized as follows. A portion of the population *ζ* ∈ [0, 1] is vaccinated and has a probability Λ ∈ [0, 1] to be infected.

### 4.3 Policy Simulation

A policy is defined on the dynamics that emerge from both the spatial and temporal sub-models, as shown in Fig. 2. First, we define the state of the model as follows.

#### Definition 1.

The Model’s state at time *t*_*i*_ is defined by the set S_*i*_ such that:

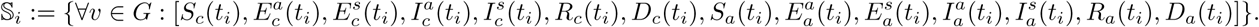

A policy ℙ is a function ℙ : {*t*_*j*_ ∈ [*t*_0_, *t*_*i*_] | 𝕊_*j*_} → Ψ such that 𝕊 ∈ Ψ that gets the states of the model from the beginning *t*_0_ and up to the current point in time *t*_*i*_ and returns a model’s state.

**Figure 2:**
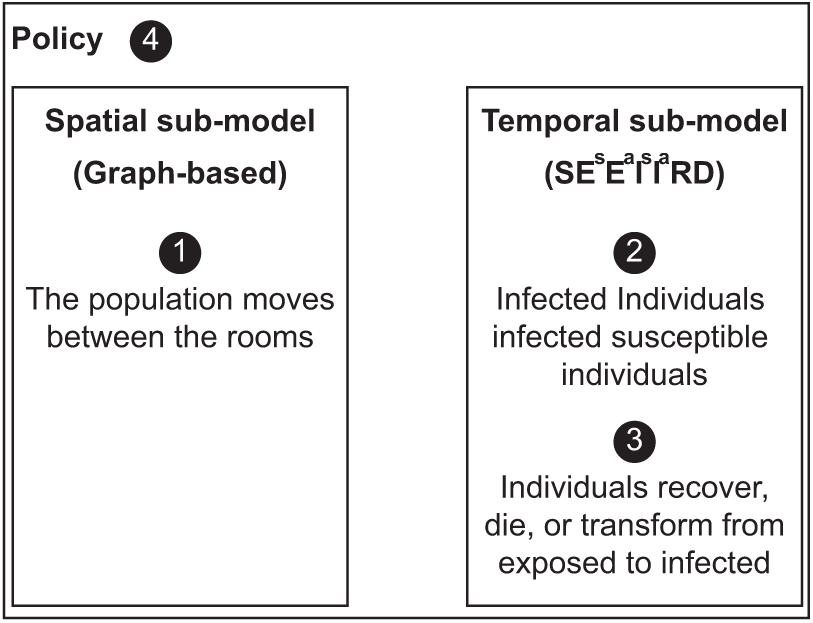
Schematic view of the model’s sub-models and their computation order for every step in time *t*_*i*_.

## 5 Results

Based on the proposed model, we examined the performance of the simulation on four building types (home, office, school, and mall). First, we gathered data regarding each building’s type topology, population, and its walk in the topology. Second, we evaluated the spread of the pandemic on all buildings, divided by type without any intervention which is defined as the *baseline* dynamics. Afterward, we examined the influence of the four proposed PIPs on each building type separately.

### 5.1 Experiment setup

We implemented the proposed model as a computer simulation. The buildings topology was obtained using maps of the building (either in physical or electronic form) or by physically mapping the building ourselves. All the buildings are located in Israel.

#### 5.1.1 Epidemiological values

We define the model’s parameters used in the following experiments. The parameters reflect the SARS-CoV-2 (COVID-19) pandemic. The parameters obtained according to biological and clinical data are presented in Table 1. The exposed to infection rate *ξ* in both asymptomatic and symptomatic and in adults and children parameters are estimated by fitting the SEEIIRD model (see Section 3.1) on the Israeli COVID-19 data [50], where *N* = 8 · 10^6^, *N*_*c*_ = 2.24 · 10^6^, and *N*_*a*_ = 5.76 · 10^6^ using the method proposed by [27], as no relevant clinical data was found.

**Table 1:** The description of the model’s parameters, values, and sources for the case of the COVID-19 pandemic.

| Parameter Definition | Symbol | Value | Source |
| --- | --- | --- | --- |
| Susceptible contacts in children which become infected due to direct disease transmission from an symptomatic/asymptomatic adult in an hour $[t^{-1}]$ | $\beta_{ca}^s, \beta_{ca}^a$ | 0.0110 | [19] |
| Susceptible contacts in adults which become infected due to direct disease transmission from symptomatic/asymptomatic children in an hour $[t^{-1}]$ | $\beta_{ac}^s, \beta_{ac}^a$ | 0.0010 | [43] |
| Susceptible contacts in children which become infected due to direct disease transmission from symptomatic/asymptomatic children in an hour $[t^{-1}]$ | $\beta_{cc}^s, \beta_{cc}^a$ | 0.0128 | [37] |
| Susceptible contacts in adults which become infected due to direct disease transmission from symptomatic/asymptomatic adult in an hour $[t^{-1}]$ | $\beta_{aa}^s, \beta_{aa}^a$ | 0.0128 | [37] |
| The probability that a child will be asymptomatic [1] | $\psi_c$ | 0.998 | [21] |
| The probability that an adult will be asymptomatic [1] | $\psi_a$ | 0.078 | [16] |
| Asymptomatic infected to recover average duration for children in hours $[t^{-1}]$ | $\gamma_c^a$ | 0.025 | [21] |
| Symptomatic infected to recover average duration for children in hours $[t^{-1}]$ | $\gamma_c^s$ | 0.02083 | [21] |
| Asymptomatic infected to recover average duration for adults in hours $[t^{-1}]$ | $\gamma_a^a$ | 0.0075 | [48] |
| Symptomatic infected to recover average duration for adults in hours $[t^{-1}]$ | $\gamma_a^s$ | 0.002975 | [48] |
| The probability an infected adult will recover from the disease [1] | $\rho_a$ | 0.942 | [33] |
| The probability an infected child will recover from the disease [1] | $\rho_c$ | 0.99 | [21] |
| The rate a symptomatic exposed child becomes infected in hours [1] | $\xi_c^s$ | 0.0018 | estimated |
| The rate a asymptomatic exposed child becomes infected in hours [1] | $\xi_a^a$ | 0.0104 | estimated |
| The rate a symptomatic exposed adult becomes infected in hours [1] | $\xi_c^s$ | 0.0056 | estimated |
| The rate a asymptomatic exposed adult becomes infected in hours [1] | $\xi_a^a$ | 0.0065 | estimated |

The model uses abstract discrete time steps. To calibrate the simulation’s abstract time step into a real one, we define each time step to be the duration of the shortest meaningful interaction between two or more individuals in a room. For example, in a school, the shortest event of the day is a 15-minute break so Δ*t* = 0.25 hours. On the other hand, if in an office each meeting or task are assumed to be in quantities of half an hour, then Δ*t* = 0.5 hours. The parameters in Table 1 are linearly scaled according to the chosen time step.

#### 5.1.2 Topology of the building and the population’s social behavior

For each building, we obtained the schematic walk of the population in one of two ways. First, the building’s operative managers (schoolmaster for the schools, chief executive officer (CEO) of the operating company of the mall, the companies’ chief operating officer (CEOs) and family members (for the *home*-type buildings) were interviewed. Second, offices had doors that open (both inside and outside) and the building’s population used a personal card that recorded each entrance and exit (per room). A log of five days was obtained and analyzed. A summary of the building types with a qualitative description of the population and topology of the building is shown in Table 2.

**Table 2:** The different building types used in the experiments with a qualitative description of the population and the topology of the building.

| Name | Population | Topology |
| --- | --- | --- |
| Home 1 | $S_a(0) = 1, S_c(0) = 2, E_a^a(0) = 1$ | $\vert V\vert = 7, \vert E\vert = 6$ |
| Home 2 | $S_a(0) = 1, E_a^a(0) = 1$ | $\vert V\vert = 7, \vert E\vert = 6$ |
| Home 3 | $S_a(0) = 2, S_c(0) = 3, E_a^a(0) = 1$ | $\vert V\vert = 6, \vert E\vert = 5$ |
| Home 4 | $S_a(0) = 2, S_c(0) = 2, E_c^a(0) = 1$ | $\vert V\vert = 7, \vert E\vert = 6$ |
| Home 5 | $S_a(0) = 2, E_c^a(0) = 1$ | $\vert V\vert = 11, \vert E\vert = 10$ |
| Office 1 | $S_a(0) = 23, E_a^a(0) = 1$ | $\vert V\vert = 10, \vert E\vert = 10$ |
| Office 2 | $S_a(0) = 46, E_a^a(0) = 1$ | $\vert V\vert = 15, \vert E\vert = 15$ |
| Office 3 | $S_a(0) = 26, E_a^a(0) = 1$ | $\vert V\vert = 10, \vert E\vert = 9$ |
| Office 4 | $S_a(0) = 11, E_a^a(0) = 1$ | $\vert V\vert = 9, \vert E\vert = 8$ |
| Office 5 | $S_a(0) = 6, I_a^a(0) = 1$ | $\vert V\vert = 5, \vert E\vert = 4$ |
| Office 6 | $S_a(0) = 17, E_a^a(0) = 1$ | $\vert V\vert = 11, \vert E\vert = 10$ |
| Office 7 | $S_a(0) = 64, E_a^a(0) = 1$ | $\vert V\vert = 25, \vert E\vert = 25$ |
| School 1 | $S_a(0) = 77, S_c(0) = 896, E_c^a(0) = 1$ | $\vert V\vert = 57, \vert E\vert = 56$ |
| School 2 | $S_a(0) = 48, S_c(0) = 450, E_a^a(0) = 1$ | $\vert V\vert = 26, \vert E\vert = 25$ |
| Mall 1 | $S_a(0) = 400, S_c(0) = 70, I_a^a(0) = 1$ | $\vert V\vert = 47, \vert E\vert = 46$ |

A schematic description of the population’s walk in each building type is provided below. The description aims to generalize the underline dynamics proposed by the representatives of the buildings shown in Table 2.

We define a *home*-type building as a house with one family. More often than not, there are both adults and children at home. While the behavior of a family changes according to its cultural, social, and economic characteristics, it is possible to draw general guidelines for the schematic behavior that a family has in its home. Specifically, both adults and children get up and prepare for the day as they spend time in the bathroom after sleeping at night in their bedroom. Afterward, part of all the individuals in the house prepare breakfast in the kitchen and eat it together in one room or separately in multiple rooms. Later, some of them spend time focusing on their tasks mostly independently in one of the rooms of the house until noon where they repeat the preparation and eating process for lunch. After lunch, some of the family members continue with their tasks until dinner where the process repeats itself for the third time. Eventually, the children repeat the morning actions as they prepare for sleep and then spend the night in their rooms. At the same time or later, the adults do the same. During the day, individuals may spend short periods in the bathroom and kitchen.

In addition, we define a *office*-type building as an office used by a single company, where all the employees of the company are adults. The employees arrive at the office during the morning at a range of times and go directly to their desks. Sometimes, the employees spend some time in the office’s kitchen if it exists. Afterward, during the day, the employees take small breaks in the kitchen or go to the bathroom. Around noon, the employees eat lunch in the kitchen or at their desks. In addition, subsets of the employees gather together at different meetings in the meeting room or in an available room in the office and sometimes even in the corridor. Finally, between afternoon and the evening employees leave the office at a range of times.

Furthermore, we define a *school* -type building as a single school which may include several physical buildings and an outdoor area in one location occupied by both adults and children. Firstly, the school’s administrative staff arrive at the school in the morning and shortly after both pupils and teachers arrive. Children have a planned program over the day where they spend the time in a classroom (either the same one or a different one according to the subject they have in the program). In between classes, pupils are allowed to move freely (and usually randomly) in the school. Teachers, on the other hand, start their day in the teachers’ lounge until the first class of the day. During a lesson, one teacher and a group of pupils are in each classroom. Teachers that do not have a class stay in the teachers’ lounge. During a break, part of the teachers are in the teachers’ lounge while the other part move randomly and supervise the pupils. Every now and then, both pupils and teachers go to the restroom. Moreover, the administrators either spend time in their offices or perform tasks in random locations in the school.

Finally, we define a *mall* -type building as an indoor shopping center with multiple shops, that serves both adults and children. One can divide the population of the mall into working-individuals such as the salespeople in the shops including the administrative staff of the mall itself and the shoppers visiting the mall. Usually, during the morning until the afternoon most of the shoppers are adults. Later on in the day, children visit the mall as well. The working-individuals are all adults. The shoppers’ sub-population can be further divided into individuals that randomly visit shops and individuals that visit a targeted list of shops to find desired items. At the end of the day, all shoppers leave the mall and only afterward the employed individuals.

## 5.2 Baseline model dynamics

Figs. 3a-3d presents the model *baseline* dynamics. The x-axis shows the time (in hours) from the beginning of the simulation and the y-axis shows the distribution of the population to *S*(*t*), *E*^*s*^(*t*), *E*^*a*^(*t*), *I*^*s*^(*t*), *I*^*a*^(*t*), *R*(*t*), and *D*(*t*). The initial condition and topology of the building are taken from Table 2 such that the results are shown as the average of the same building type for *n* = 100 repetitions each.

**Figure 3:**
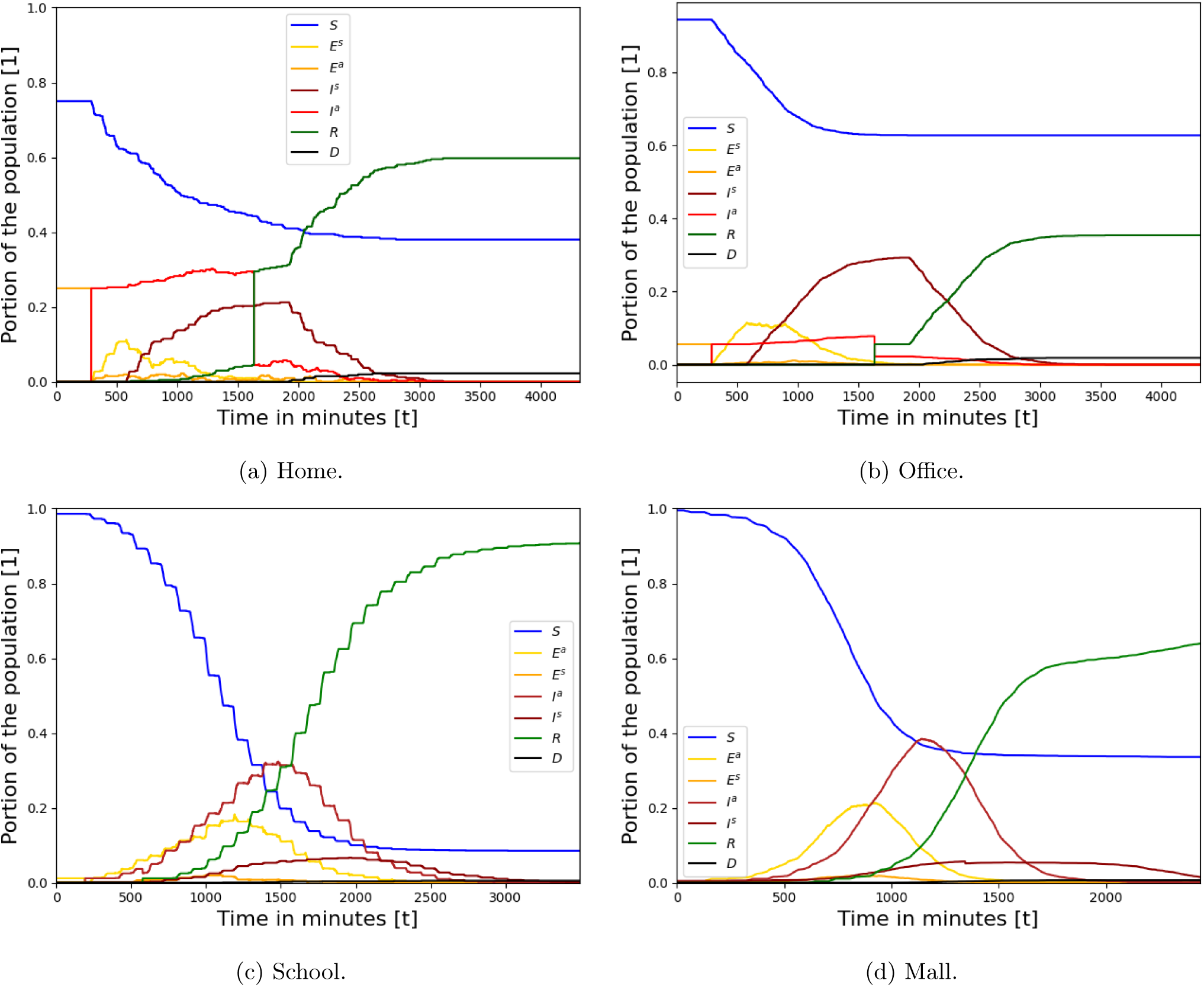
Baseline dynamics of the epidemiological spread in the population, divided by the building types. The results are the mean of *n* = 100 repeats. The epidemiological parameters are taken from Table 1 and the social and topological parameters are taken from Section 5.1.2 and Table 2, respectively.

### 5.3 Sensitivity Analysis

Figs. 4-7 present the model sensitivity for each one of the PIPs (see Section 4) for each building type. The figures are divided into four plots as follows: First, the ISII policy sensitivity graphs, where the x-axis shows the rate of tests (*τ*) and the y-axis shows the portion of rooms (*σ*) that are included in each test. Second, the SD policy sensitivity graphs, where the x-axis shows the probability (*ξ*) that an individual overrides its original walk dynamics with one optimization of social distancing and the y-axis shows the number of infected individuals from the population. Third, the MW policy sensitivity graphs, where the x-axis shows the average quality of mask-wearing in reducing infection rate (*α*) and the y-axis shows the portion of the population that wears masks (Γ). Finally, the vaccination policy sensitivity graphs, where the x-axis shows the efficiency of vaccination in reducing infection (Λ) and the y-axis shows the portion of the population that is vaccinated (*ζ*). In all plots of PIPs with two parameters, the color indicates the portion of infected individuals from the population.

**Figure 4:**
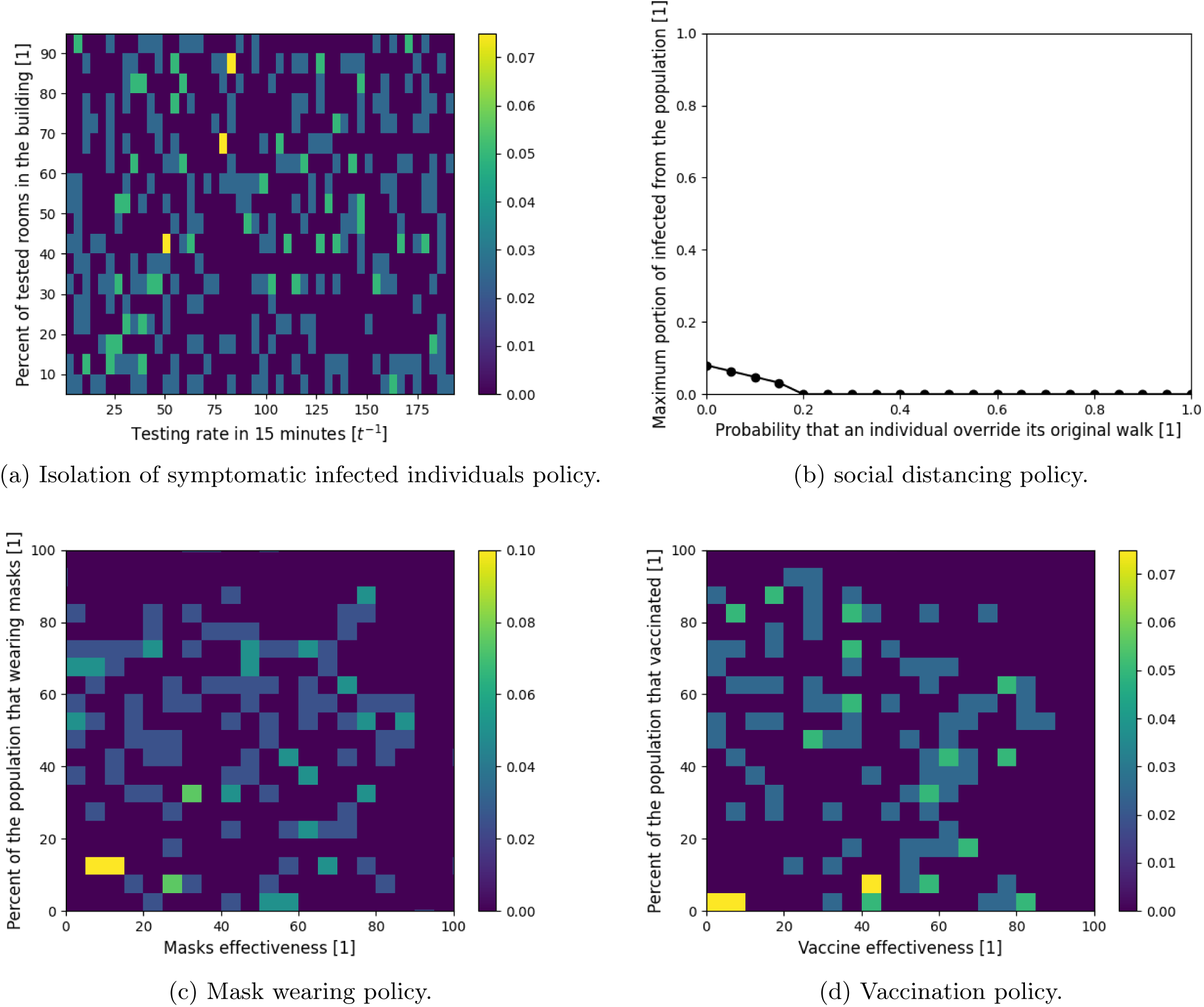
Sensitivity analysis of the parameter space of each PIP for the *home*-type building. The results are the mean of *n* = 30 repeats for each of the houses. The epidemiological parameters are taken from Table 1 and the social and topological parameters are taken from Section 5.1.2 and Table 2, respectively.

**Figure 5:**
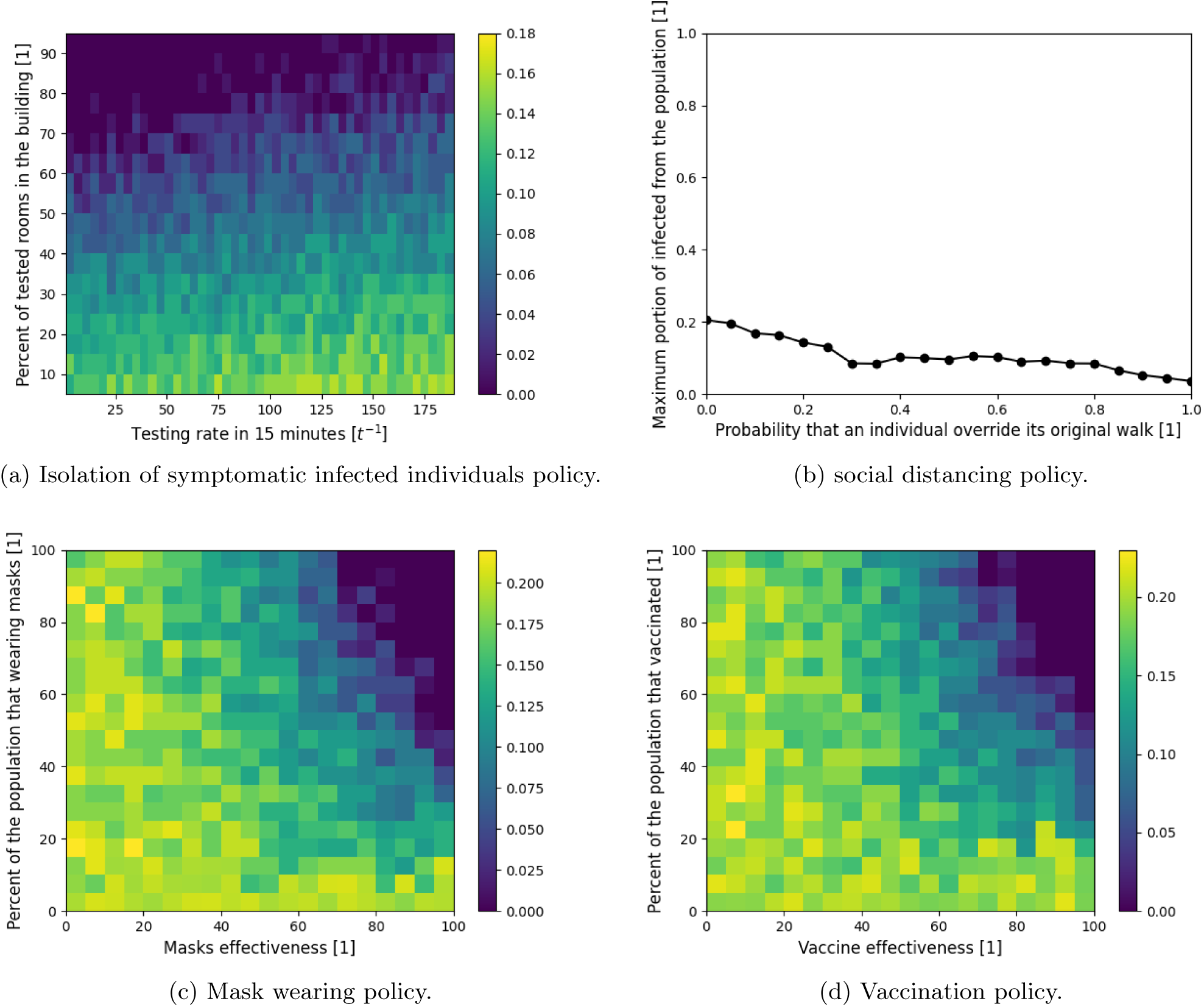
Sensitivity analysis of the parameter space of each PIP for the *office*-type building. The results are the mean of *n* = 30 repeats for each of the houses. The epidemiological parameters are taken from Table 1 and the social and topological parameters are taken from Section 5.1.2 and Table 2, respectively.

**Figure 6:**
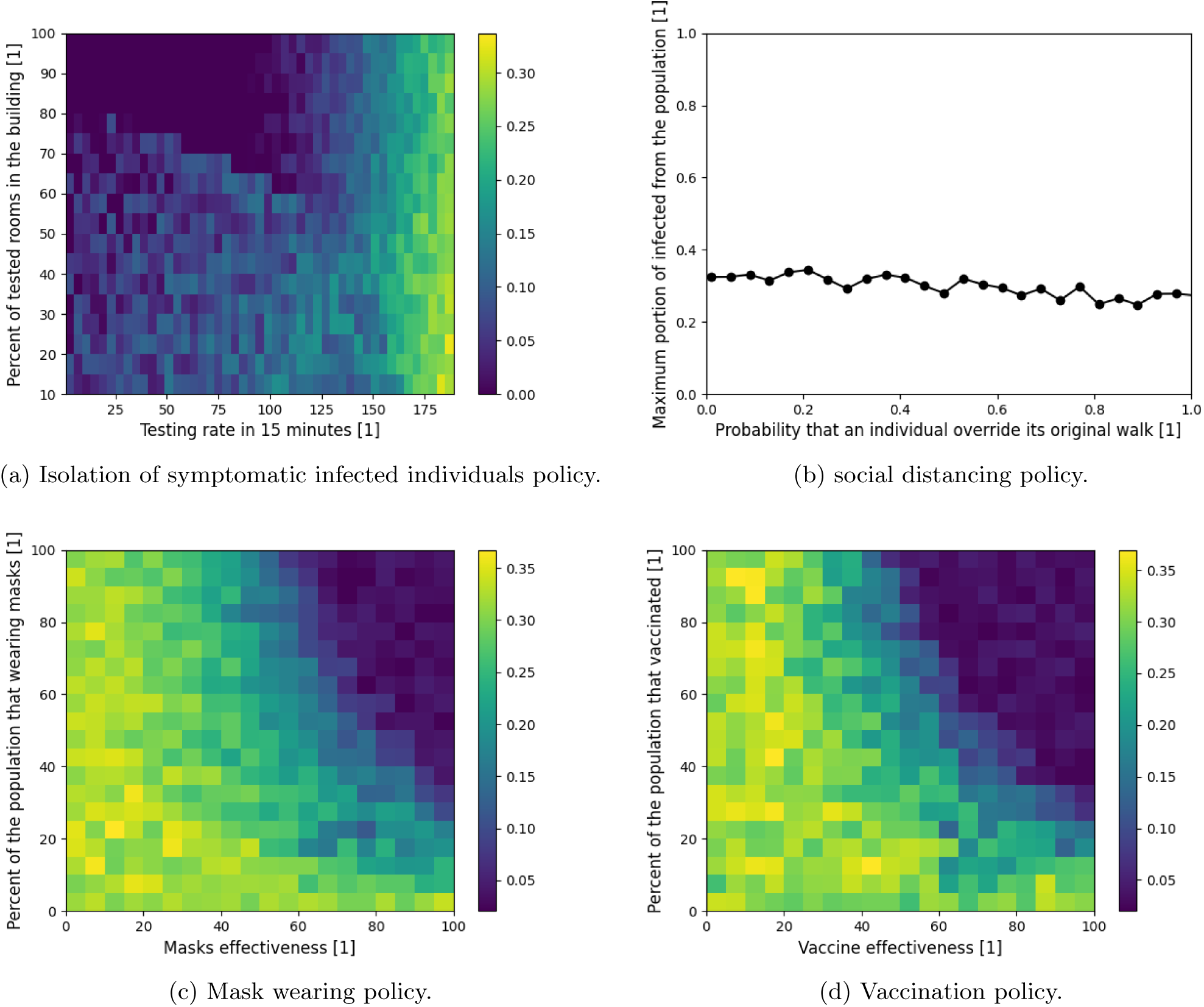
Sensitivity analysis of the parameter space of each PIP for the *school* -type building. The results are the mean of *n* = 30 repeats for each of the houses. The epidemiological parameters are taken from Table 1 and the social and topological parameters are taken from Section 5.1.2 and Table 2, respectively.

**Figure 7:**
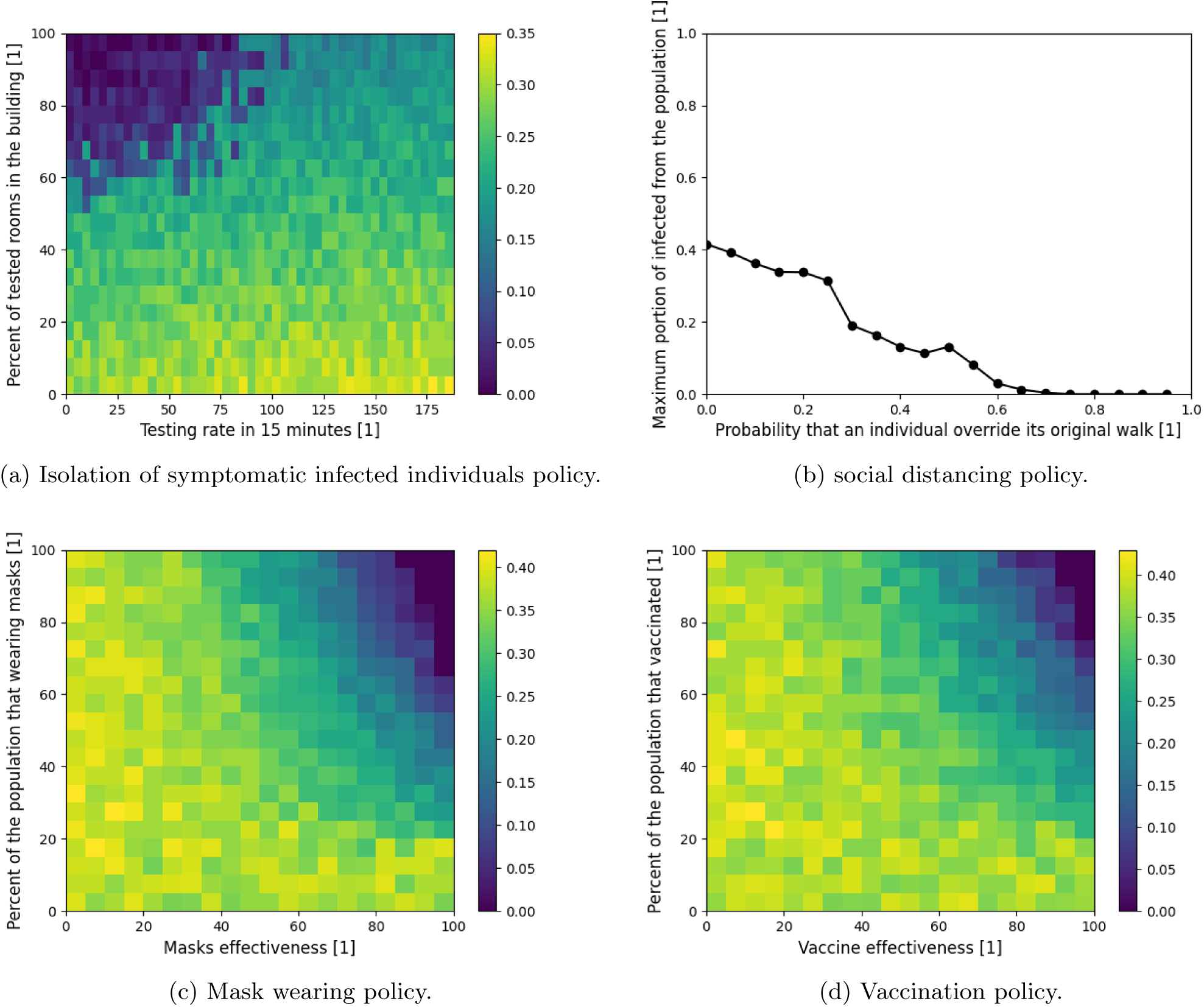
Sensitivity analysis of the parameter space of each PIP for the *mall* -type building. The results are the mean of *n* = 30 repeats for each of the houses. The epidemiological parameters are taken from Table 1 and the social and topological parameters are taken from Section 5.1.2 and Table 2, respectively.

In order to evaluate the average influence of each parameter of the PIPs on the pandemic spread in the context of each building type, we fitted the results from the simulation. The fitting function is calculated using the least mean square (LMS) method [7]. In order to use the LMS method, one needs to define the family function approximating a function. The one dimensional and two dimensional linear family function (e.g., *f* (*x, y*) = *p*_1_ + *p*_2_*x* + *p*_3_*y*) are chosen to obtain the linear influence of each parameter - in the form of the coefficient of the respected parameter in the fitted parameter. The results are shown in Table 3.

**Table 3:** Linear fit of the parameter space of each PIP for each building type. The coefficients are rounded up to three digits after the decimal point. ^*^ Fitted on *χ* ∈ [0, 0.2] and for *chi* ∈ [0.2, 1], *I* = 0 (*R*^2^ = 1).

|  | Home | Office | School | Mall |
| --- | --- | --- | --- | --- |
| **ISII** | Not converged | $-0.002\tau - 0.006\sigma + 0.777$ ($R^2 = 0.73$) | $0.003\tau - 0.003\sigma + 0.220$ ($R^2 = 0.86$) | $0.001\tau - 0.002\sigma + 0.304$ ($R^2 = 0.95$) |
| **SD** | $-0.296\chi + 0.064$ ($R^2 = 0.96$)* | $-0.169\chi + 0.198$ ($R^2 = 0.81$) | $-0.045\chi + 0.261$ ($R^2 = 0.52$) | $-0.473\chi + 0.325$ ($R^2 = 0.97$) |
| **MW** | Not converged | $-0.001\alpha - 0.001\Gamma + 0.2261$ ($R^2 = 0.73$) | $-0.001\alpha - 0.002\Gamma + 0.387$ ($R^2 = 0.57$) | $-0.001\alpha - 0.002\Gamma + 0.489$ ($R^2 = 0.78$) |
| **Vaccination** | Not converged | $-0.001\zeta - 0.001\Lambda + 0.229$ ($R^2 = 0.62$) | $-0.001\zeta - 0.002\Lambda + 0.407$ ($R^2 = 0.80$) | $-0.001\zeta - 0.001\Lambda + 0.401$ ($R^2 = 0.32$) |

## 6 Discussion

In future pandemics, policymakers will be able to use our model to investigate the consequences of several PIPs on the pandemic spread. The model has seven extensions to the traditional SIR model: separation of the population into two age groups (children and adults); separation of the infected group into asymptomatic and symptomatic infected groups; introduction of a dead group (*D*); introduction of asymptomatic and symptomatic exposed groups, including graph-based spatial dynamics; use of the *Wells-Riley* [41] in-room pandemic spread; and heterogeneous walks.

Using these extensions, the model shows that pandemic spread highly differs in homes, offices, schools, and malls as shown in Fig. 3. The model shows the pandemic spread in homes is more chaotic than the other building types, as shown in Fig. 3a, which can be associated with the low density of the population (average number of individuals in a room) which is 0.53 compared to 4.06, 17.75, 10.02 in the office, school, and mall, respectively. On the other hand, due to low density, a large portion (38% on average) is kept uninfected even without any PIPs. The (high-tech) office is the most pandemic spread resistant, as shown in Fig. 3b, and can be explained by the fact that individuals are staying in their rooms most of the time and therefore produce only a local spread. On the other hand, the model shows that schools are the building type with the highest infection rate (with around 90% infection), as shown in Fig. 3c. There are several reasons for this. First, pupils spend long periods in the classrooms which results in infection between classmates, as shown by the drops in the susceptible (blue) population size. Second, during breaks, the pupils move around the building randomly which operates as a bridge of infection between classrooms. Third, infected adults (teachers, maintenance employees, etc.) operate as an infection vector as they move between large groups of pupils. In the case of a mall, the pandemic spread is similar to the standard two-age SIR [9] model with one location. This can be explained by the large portion of the population that performs a somewhat random walk which is known to approximate the temporal SIR model [17]. However, some portion of the population performs a targeted buying which makes it less likely to be infected and therefore not (almost) the entire population being infected at some point.

Based on the proposed model, we evaluated four PIPS by the portion of the total number of infected individuals from the population. The four PIPs are isolation of symptomatic infected individuals, social distancing, mask-wearing, and vaccination. The model shows that all four PIPs are random for the home building type, as shown in Figs. (4a, 4c, and 4d). This behavior is associated with the low density of the population together with the small population size which results in large differences even by changes in a single individual’s state. As a result, the social distancing PIP is very useful, as individuals can find empty rooms to stay in and thereby avoid infections, as shown in Fig. 4b.

Furthermore, we evaluated four pandemic intervention policies (PIPs) - isolated symptomatic infected individuals (ISII), social distancing (SD), mask wearing (MW), and vaccination. Each PIP is evaluated on each of the four building types, showing the influence of the pandemic spread on the range of the parameters that define each PIP as shown in Figs. 4-7. Linear regression has been performed on each simulation result for a specific PIP and building type to obtain the linear influence of each PIPs’ parameters on the pandemic spread for each building type as shown in Table 3.

Similar to the baseline dynamics of the *home*-type building, the ISII, MV, and vaccination PIPs do not have a clear behavior that can be associated with small population size and low density together. On the other hand, the SD PIP is able to reduce the pandemic spread such that on average if 20% of the time individuals were to avoid being in the same room without other individuals, the pandemic would spread after the first infected individual which exploits the low density. As a result, the SD PIP is the most suitable for homes while other PIPs (even vaccination) have a chaotic behavior which is an issue for policymakers.

For the *office, school*, and *mall* building types, the MV and vaccination PIPs have similar average behavior as shown in Table 3. Indeed, as temporal PIP are less sensitive to spatial dynamics (e.g., individuals’ walks in the building). In addition, by definition, the MW and vaccination are similar, the results show that indeed over several building types the influence of these PIPs on the pandemic spread is equal while the baseline (the free coefficient) differs between building types. On the other hand, the ISII PIP is a spatial-based PIP and indeed differs between the building types such that in the *office* the testing rate is more important than the coverage of testing, in the *school* both have the same influence on the pandemic spread, and in the *mall*, it is the opposite to the *office*, as shown in Table 3. Similarly, the SD PIP differ between the building types, showing a quick decrease to zero in the *mall* (70%) and a slower one in the *office* (15% from the case without SD) building types while less than 20% influence in the case of a *school* building type. A possible reason for the inefficiency of the SD in the school is the relatively high density compared to the *office* and *mall* building types which makes the SD less effective in practice.

Therefore, while there is a similarity between the temporal-based PIP between building types, spatial-based PIP highly differs in their effectiveness to control the pandemic spread.

One limitation of the proposed agent-based simulator is that it is linear (*O*(|*P*|)) to the size of the population (*P*) which is significantly worse than solving the ODE at each point in time (which is independent of the size of the population and therefore computes in *O*(1)). Therefore, for large size populations such as in the case of cities, countries, or the entire world’s population the computation increases. However, most of the computation steps in the simulations are independent in the scope of a single individual in the population which allows relatively easy parallelization of the computation which can reduce the computing time to make it feasible even for large populations.

## 7 Conclusion

The model developed in this study allows us to examine the impact of non-pharmaceutical and pharmaceutical PIPs on the course of a pandemic spread in the context of a single building. The model is implemented for the COVID-19 outbreak with data of buildings from Israel. It extends the traditional SIR model by introducing asymptomatic and symptomatic exposed groups and a dead group, splitting the infected group into asymptomatic and symptomatic infected two age-groups, including graph-based locations where individuals can be present during the day, and introducing a heterogeneous walk on the graph for each individual in the population. These spatio-temporal interactions allow us to explore the reciprocal effects of both spatial and temporal PIP on the spread of the pandemic in different buildings and social contexts such as the effect of mask-wearing in school as shown in Fig. 6c. The inclusion of these interactions improves the accuracy of the model’s forecasts and allows for multidimensional analysis of the impact of PIPs and the dynamics of the crisis.

Our results indicate that policymakers need to take into consideration the unique social properties that individuals in the population have in different buildings (in which they spend most of their time) to find the optimal PIP as differential mask wearing among different age groups or social roles (such as teachers and pupils in schools), or varying symptomatic infection testing, that significantly improve policy trade-offs, enabling considerable reductions in the pandemic spread and excess deaths.

The results in this study were obtained given the values in Table 1, buildings data from Table 2, and walks description from Section 5.1.2 which can vary significantly across countries and time as it depend on several hyper-parameters such as the local culture, architectural style, and population density. However, the proposed model does not take into consideration the movement of individuals between buildings during the day and the differences between the days of the week (such as weekends and working days). Assuming such dynamics, the results shown in Fig. 3 could be significantly altered. Therefore, we intend to include these dynamics in future studies.

## Data Availability

The buildings raw topological data is not available due to security reasons but the graph representation that has been used in the simulation is available upon request from the authors.

## References

[1] O. Aglar, A. Baxter, P. Keskinocak, J. Asplund, and N. Serban. “Homebound by COVID19: The Benefits and Consequences of Non-pharmaceutical Intervention Strategies”. In: Research Square (2020).

[2] L. J. S. Allen. “Some discrete-time SI, SIR, and SIS epidemic models”. In: Mathematical Biosciences 124.1 (1994), pp. 83–105.

[3] Lydia Aslanidou, Bram Trachet, Philippe Reymond, Rodrigo A. Fraga-Silva, Patrick Segersm, and Nikolas Stergiopulos. “A 1D Model of the Arterial Circulation in Mice”. In: Journal Biomechanics (2016).

[4] B. F. Balvedi, E. Ghisi, and R. Lamberts. “A review of occupant behaviour in residential buildings”. In: Energy Buildings 174 (2018), pp. 495–505.

[5] O. Barnea, R. Yaari, G. Katriel, and L. Stone. “Modeling seasonal influenze in Israel”. In: Mathematical Biosciences and Engineering 8 (2011), pp. 561–573.

[6] F. Bernaridini and M. Ghorghe. “Population P Systems”. In: Journal of Universal Computer Science 10 (2004), pp. 509–539.

[7] A. Bjorck. “Numerical Methods for Least Squares Problems”. In: Society for Industrial and Applied Math-matics 5 (1996), pp. 497–513.

[8] A. Brodeur, D. Gray, A. Islam, and S. Bhuiyan. “A Literature Review of the Economics of COVID-19”. In: IZA Discussion Paper No. 13411, Available at SSRN: https://ssrn.com/abstract=3636640 (2020).

[9] S. Bunimovich-Mendrazitsky and L. Stone. “Modeling polio as a disease of development”. In: Journal of Theoretical Biology 237 (2005), pp. 302–315.

[10] Andrea Alberto Conti. “Historical and methodological highlights of quarantine measures: from ancient plague epidemics to current coronavirus disease (COVID-19) pandemic”. In: Acta bio-medica : Atenei Parmensis 91.2 (2020), pp. 226–229.

[11] J-C. Cortés, S. K. El-Labany, A. Navarro-Quiles, M. M. Selim, and H. Slama. “A comprehensive probabilistic analysis of approximate SIR-type epidemiological models via full randomized discrete-time Markov chain formulation with applications”. In: Mathematical Methods in the Applied Sciences 43.14 (2020), pp. 8204–8222.

[12] S. F. Darabi and C. Scoglio. “Epidemic spread in human networks.” In: 50th IEEE Conference on Decision and Control and European Control Conference. 2011, pp. 3008–3013.

[13] L. Di Domenico, G. Pullano, C. E. Sabbatini, P. Y. Bo Elle, and V. Colizza. “Impact of lockdown on COVID-19 epidemic in Ile-de-France and possible exit strategies”. In: BMC Medicine 18 (2020).

[14] B. L. Diffey. “An overview analysis of the time people spend outdoors”. In: British Journal of Dermatology 164 (2010), pp. 848–854.

[15] G. Hamra, R. MacLehose, and D. Richardson. “Markov chain Monte Carlo: an introduction for epidemiologists”. In: Int J Epidemiology 42 (2013), pp. 627–634.

[16] J. He, Y. Guo, R. Mao, and J. Zhang. “Proportion of asymptomatic coronavirus disease 2019: A systematic review and meta-analysis”. In: J Med Virol (2020), pp. 1–11.

[17] H-F. Huo, Q. Yang, and H. Xiang. “Dynamics of an edge-based SEIR model for sexually transmitted diseases”. In: Mathematical Biosciences and Engineering 17 (2019), pp. 669–699.

[18] B. Ivorra, M. R. Ferrandez, M. Vela-Perez, and A. M. Ramos. “Mathematical modeling of the spread of the coronavirus disease 2019 (COVID-19) taking into account the undetected infections. The case of China”. In: Communications in nonlinear science and numerical simulation 88 (2020), p. 105303.

[19] Cai Jiehao, Xu Jin, Lin Daojiong, Yang Zhi, Xu Lei, Qu Zhenghai, Zhang Yuehua, Zhang Hua, Jia Ran, Liu Pengcheng, Wang Xiangshi, Ge Yanling, Xia Aimei, Tian He, Chang Hailing, Wang Chuning, Li Jingjing, Wang Jianshe, and Zeng Mei. “A Case Series of Children With 2019 Novel Coronavirus Infection: Clinical and Epidemiological Features”. In: Clinical Infectious Diseases (2020).

[20] S. R. Keilich, J. M. Bartley, and L. Haynes. “Diminished immune responses with aging predispose older adults to common and uncommon influenza complications”. In: Cellular Immunology 345 (2019), p. 103992.

[21] A. A. Kelvin and S. Helperin. “COVID-19 in children: the link in the transmission chain”. In: Lancet 20 (2020), pp. 633–634.

[22] W. O. Kermack and A. G. McKendrick. “A contribution to the mathematical theory of epidemics”. In: Proceedings of the Royal Society 115 (1927), pp. 700–721.

[23] J. C. King, J. J. Stoddard, M. J. Gaglani, K. A. More, L. Magder, E. McClure, J. D. Rubin, J. A. Englund, and K. Neuzil. “Effectiveness of School-Based Influenza Vaccination”. In: The New England Journal of Medicine 355 (2006), pp. 2523–2532.

[24] D. Kingsley. The Urbanization of the Human Population. Routledge, 2015.

[25] S. A. Lauer, K. H. Grantz, Q. Bi, F. K. Jones, Q. Zheng, H. R. Meredith, A. S. Azman, N. G. Reich, and J. Lessier. “The Incubation Period of Coronavirus Disease 2019 (COVID-19) From Publicly Reported Confirmed Cases: Estimation and Application”. In: Annals of Internal Medicine 172.9 (2020), pp. 577– 582.

[26] T. Lazebnik and S. Bunimovich-Mendrazitsky. “The signature features of COVID-19 pandemic in a hybrid mathematical model - implications for optimal work-school lockdown policy”. In: advanced theory and simulations (2021).

[27] T. Lazebnik, L. Shami, and S. Bunimovich-Mendrazitsky. “Spatio-Temporal Influence of Non-Pharmaceutical Interventions Policies on Pandemic Dynamics and the Economy: The Case of COVID-19”. In: Research Economics (2021).

[28] Joshua Lederberg. “Medical Science, Infectious Disease, and the Unity of Humankind”. In: JAMA 260.5 (1988), pp. 684–685.

[29] T. Li, Y. Liu, Li M., X. Qian, and S. Y. Dai. “Mask or no mask for COVID-19: A public health and market study”. In: PLoS ONE 15 (2020), e0237691.

[30] C. M. Macal. “To agent-based simulation from System Dynamics”. In: Proceedings of the 2010 Winter Simulation Conference. 2010, pp. 371–382.

[31] Frantisek Marsik, Svetlana Prevorovska, Zdenek Broz, and Vitezslav Stembera. “Numerical Model of the Human Cardiovascular System–Korotkoff Sound Simulation”. In: Cardiovascular Engineering: and International Journal 4 (2004).

[32] S. Masud, V. Torraca, and A. H. Meijer. “Chapter Eight - Modeling Infectious Diseases in the Context of a Developing Immune System”. In: Zebrafish at the Interface of Development and Disease Research. Ed. by Kirsten C. Sadler. Vol. 124. Current Topics in Developmental Biology. Academic Press, 2017, pp. 277–329.

[33] M. R. Mehra, S. S. Desai, S. Kuy, T. D. Henry, and A. N. Patel. “Cardiovascular Disease, Drug Therapy, and Mortality in COVID-19”. In: The New England Journal of Medicine 382 (2020), e102.

[34] J. C. Miller. “Mathematical models of SIR disease spread with combined non-sexual and sexual transmission routes”. In: Infectious Disease Modelling 2 (2017), pp. 35–55.

[35] S. Moore, E. M. Hill, M. J. Tildesley, L. Dyson, and P. M. J. Keeling. “Vaccination and non-pharmaceutical interventions for COVID-19: a mathematical modelling study”. In: The Lancet (2021).

[36] L. Nesteruk. “Statistics-based Predictions of Coronavirus Epidemic Spreading in Mainland China”. In: Innov Biosyst Bioeng 8 (2020), pp. 13–18.

[37] H. Nishiura and T. Kobayashi. “Estimation of the asymptomatic ratio of novel coronavirus infections (COVID-19)”. In: International Journal of Infections Diseases 94 (2020), pp. 154–155.

[38] C. J. Noakes, C. B. Beggs, P. A. Sleigh, and K. G. Kerr. “Modelling the transmission of airborne infections in enclosed spaces”. In: Epidemiol. Infect. 134 (2006), pp. 1082–1091.

[39] K. O’Dowd, K. M. Nair, P. Forouzanadeh, S. Mathew, J. Grant, R. Moran, J. Bartlett, J. Bird, and S. C. Pillai. “Face Masks and Respirators in the Fight Against the COVID-19 Pandemic: A Review of Current Materials, Advances and Future Perspectives”. In: Materials 13 (2020), p. 3363.

[40] G. Paun. “Computing with Membranes”. In: Journal of Computer and System Sciences 61 (2000), pp. 108–143.

[41] E.C. Riley, G. Murphy, and R. L. Riley. “Airborne spread of measles in a suburban elementary school”. In: Journal of Epidemiology 107 (1978), pp. 421–432.

[42] L. Ronald. “The Outlook for Population Growth”. In: Science 333.6042 (2011), pp. 569–573.

[43] Jiatong She, Lanqin Liu, and Wenjun Liu. “COVID-19 epidemic: Disease characteristics in children”. In: Journal of medical virology (2020).

[44] Eurosurveillance Editorial Team. “Note from the editors: World Health Organization declares novel coronavirus (2019-nCoV) sixth public health emergency of international concern”. In: Euro Surveill 25 (2020), 200131e.

[45] A. R. Tuite, D. N. Fisman, and A. L. Greer. “Mathematical modelling of COVID-19 transmission and mitigation strategies in the population of Ontario, Canada”. In: CMAJ 192 (2020), E497–E505.

[46] A. Viguerie, G. Lorenzo, F. Auricchio, D. Baroli, T. J. R. Hughes, A. Patton, A. Reali, T. E. Yankeelov, and A. Veneziani. “Simulating the spread of COVID-19 via a spatially- resolved susceptible– exposed–infected– recovered– deceased (SEIRD) model with heterogeneous diffusion”. In: Applied Mathematics Letters 111 (2020).

[47] Victor Virlogeux, Ming Li, Tim K. Tsang, Luzhao Feng, Vicky J. Fang, Hui Jiang, Peng Wu, Jiandong Zheng, Eric H. Y. Lau, Yu Cao, Ying Qin, Qiaohong Liao, Hongjie Yu, and Benjamin J. Cowling. “Estimating the Distribution of the Incubation Periods of Human Avian Influenza A(H7N9) Virus Infections”. In: American Journal of Epidemiology 182.8 (2015), pp. 723–729.

[48] Voinsky, G. Baristaite, and D. Gurwitz. “Effects of age and sex on recovery from COVID-19: Analysis of 5769 Israeli patients”. In: The Journal of infection 81 (2020), pp. 102–103.

[49] X. Wang, Z. Wang, and H. Shen. “Dynamical analysis of a discrete-time SIS epidemic model oncomplex networks”. In: Applied Mathematics Letters 94 (2019), pp. 292–299.

[50] WHO. WHO Coronavirus Disease (COVID-19) Dashboard. url: https://covid19.who.int/. (accessed: 25.05.2021).

[51] T. Wu, C. Perrings, A. Kinzig, J. P. Collins, B. A. Minteer, and P. Daszak. “Economic growth, urbanization, globalization, and the risks of emerging infectious diseases in China: A review”. In: Ambio 46.1 (2017), pp. 18–29.

[52] W. Yang, D. Zhang, L. Peng, C. Zhuge, and L. Liu. “Rational evaluation of various epidemic models based on the COVID-19 data of China”. In: medRxiv 344 (2020).

[53] S. Zhao, L. Stone, D. Gao, S. S. Musa, M. K. C. Chong, D. He, and M. H. Wang. “Imitation dynamics in the mitigation of the novel coronavirus disease (COVID-19) outbreak in Wuhan, China from 2019 to 2020”. In: Annals of Transnational Medicine 8 (2020).

