## Appendix for "Comparison of Pandemic Intervention Policies in Several Building Types Using Heterogeneous Population Model"

Teddy Lazebnik Ariel Alexi
Computer Science Department, Bar Ilan University, Israel

### 1 Epidemiological (Temporal) sub-model

The epidemiological dynamics is described in Eqs. (S1-S14).

In Eq. (S1), $\frac{dS_c(t)}{dt}$ is the dynamic amount of susceptible children over time. It is affected by the following four terms: 1) each symptomatic infected child infects susceptible children at a rate $\beta^s_{cc}$; 2) each asymptomatic infected child infects susceptible children at a rate $\beta^a_{cc}$; 3) each infected symptomatic adult infects the susceptible children at a rate $\beta^s_{ca}$; 4) each infected asymptomatic adult infects the susceptible children at a rate $\beta^a_{ca}$. $N_c$ is the size of the children population and used to take all variables as fixed proportions of the population $N$.

$$\frac{dS_c(t)}{dt} = -\frac{\beta^s_{cc} I^s_c(t) + \beta^a_{cc} I^a_c(t) + \beta^s_{ca} I^s_a(t) + \beta^a_{ca} I^a_a(t)}{N_c} S_c(t). \tag{S1}$$

In Eq. (S2), $\frac{dS_a(t)}{dt}$ is the dynamic amount of susceptible adult over time. It is affected by the following four terms: 1) each symptomatic infected child infects the susceptible adult at a rate $\beta^s_{ac}$; 2) each asymptomatic infected child infects the susceptible adult at a rate $\beta^a_{ac}$; 3) each symptomatic infected adult infects a susceptible adult at a rate $\beta^s_{aa}$; 4) each asymptomatic infected adult infects a susceptible adult at a rate $\beta^a_{aa}$. $N_a$ is the size of the adult population and used to take all variables as fixed proportions of the population $N$.

$$\frac{dS_a(t)}{dt} = -\frac{\beta^s_{ac} I^s_c(t) + \beta^a_{ac} I^a_c(t) + \beta^s_{aa} I^s_a(t) + \beta^a_{aa} I^a_a(t)}{N_a} S_a(t). \tag{S2}$$

In Eq. (S3), $\frac{dE^s_c(t)}{dt}$ is the dynamic amount of symptomatic exposed children over time. It is affected by the following five terms: 1) each symptomatic infected child infects susceptible children at a rate $\beta^s_{cc}$; 2) each asymptomatic infected child infects susceptible children at a rate $\beta^a_{cc}$; 3) each infected symptomatic adult infects the susceptible children at a rate $\beta^s_{ca}$; 4) each infected asymptomatic adult infects the susceptible children at a rate $\beta^a_{ca}$; 5) each symptomatic exposed child is transformed into an symptomatic infected child at a rate $\xi^s_c$. The infection terms (1-4) are multiplied by the probability a child will be symptomatic $1 - \psi_c$.

$$\frac{dE^s_c(t)}{dt} = (1 - \psi_c)\frac{\beta^s_{cc} I^s_c(t) + \beta^a_{cc} I^a_c(t) + \beta^s_{ca} I^s_a(t) + \beta^a_{ca} I^a_a(t)}{N_c} S_c(t) - \xi^s_c E^s_c(t). \tag{S3}$$

In Eq. (S4), $\frac{dE^a_c(t)}{dt}$ is the dynamic amount of asymptomatic exposed children over time. It is affected by the following five terms: 1) each symptomatic infected child infects susceptible children at a rate $\beta^s_{cc}$; 2) each asymptomatic infected child infects susceptible children at a rate $\beta^a_{cc}$; 3) each infected symptomatic adult infects the susceptible children at a rate $\beta^s_{ca}$; 4) each infected asymptomatic adult infects the susceptible children at a rate $\beta^a_{ca}$; 5) each asymptomatic exposed child is transformed into an symptomatic infected child at a rate $\xi^a_c$. The infection terms (1-4) are multiplied by the probability a child will be asymptomatic $\psi_c$.

$$\frac{dE^a_c(t)}{dt} = \psi_c \frac{\beta^s_{cc} I^s_c(t) + \beta^a_{cc} I^a_c(t) + \beta^s_{ca} I^s_a(t) + \beta^a_{ca} I^a_a(t)}{N_c} S_c(t) - \xi^a_c E^a_c(t). \tag{S4}$$

In Eq. (S5), $\frac{dE^s_a(t)}{dt}$ is the dynamic amount of symptomatic exposed adult over time. It is affected by the following five terms: 1) each symptomatic infected child infects the susceptible adult at a rate $\beta^s_{ac}$; 2) each asymptomatic infected child infects the susceptible adult at a rate $\beta^a_{ac}$; 3) each symptomatic infected adult infects a susceptible adult at a rate $\beta^s_{aa}$; 4) each asymptomatic infected adult infects a susceptible adult at a rate $\beta_{aa}^{a}$; 5) each symptomatic exposed adult is transformed into an symptomatic infected adult at a rate $\xi_a^s$. The infection terms (1-4) are multiplied by the probability an adult will be symptomatic $1 - \psi_a$.

$$\frac{dE_a^s(t)}{dt} = (1 - \psi_a)\frac{\beta_{ac}^s I_c^s(t) + \beta_{ac}^a I_c^a(t) + \beta_{aa}^s I_a^s(t) + \beta_{aa}^a I_a^a(t)}{N_a} S_a(t) - \xi_a^s E_a^s(t)). \tag{S5}$$

In Eq. (S6), $\frac{dE_a^a(t)}{dt}$ is the dynamic amount of asymptomatic exposed adult over time. It is affected by the following five terms: 1) each symptomatic infected child infects the susceptible adult at a rate $\beta_{ac}^s$; 2) each asymptomatic infected child infects the susceptible adult at a rate $\beta_{ac}^a$; 3) each symptomatic infected adult infects a susceptible adult at a rate $\beta_{aa}^s$; 4) each asymptomatic infected adult infects a susceptible adult at a rate $\beta_{aa}^a$; 5) each symptomatic exposed adult is transformed into an symptomatic infected adult at a rate $\xi_a^a$. The infection terms (1-4) are multiplied by the probability an adult will be asymptomatic $\psi_a$.

$$\frac{dE_a^a(t)}{dt} = \psi_a \frac{\beta_{ac}^s I_c^s(t) + \beta_{ac}^a I_c^a(t) + \beta_{aa}^s I_a^s(t) + \beta_{aa}^a I_a^a(t)}{N_a} S_a(t) - \xi_a^a E_a^a(t). \tag{S6}$$

In Eq. (S7), $\frac{dI_c^s(t)}{dt}$ is the dynamic amount of symptomatic infected children over time. It is affected by the following two terms: 1) individuals recover or die from the disease after period $\gamma_c^s$; 2) each symptomatic exposed child is transformed into an symptomatic infected child at a rate $\xi_c^s$.

$$\frac{dI_c^s(t)}{dt} = \xi_c^s E_c^s(t) - \gamma_c^s I_c^s(t). \tag{S7}$$

In Eq. (S8), $\frac{dI_c^a(t)}{dt}$ is the dynamic amount of asymptomatic infected children over time. It is affected by the following two terms: 1) individuals recover from the disease after period $\gamma_c^a$; 2) each asymptomatic exposed child is transformed into an asymptomatic infected child at a rate $\xi_c^a$.

$$\frac{dI_c^a(t)}{dt} = \xi_c^a E_c^a(t) - \gamma_c^a I_c^a(t). \tag{S8}$$

In Eq. (S9), $\frac{dI_a^s(t)}{dt}$ is the dynamic amount of symptomatic infected adult over time. It is affected by the following two terms: 1) individuals recover or die from the disease after period $\gamma_a^s$; 2) each symptomatic exposed adult is transformed into an symptomatic infected adult at a rate $\xi_a^s$.

$$\frac{dI_a^s(t)}{dt} = \xi_a^s E_a^s(t) - \gamma_a^s I_a^s(t). \tag{S9}$$

In Eq. (S10), $\frac{dI_a^a(t)}{dt}$ is the dynamic amount of asymptomatic infected adult over time. It is affected by the following two terms: 1) individuals recover from the disease after period $\gamma_a^a$; 2) each asymptomatic exposed adult is transformed into an asymptomatic infected adult at a rate $\xi_a^a$.

$$\frac{dI_a^a(t)}{dt} = \xi_a^a E_a^a(t) - \gamma_a^a I_a^a(t). \tag{S10}$$

In Eq. (S11), $\frac{dR_c(t)}{dt}$ is the dynamic amount of recovered children over time. It is affected by the following two terms: 1) in each point, a portion of the symptomatic infected children recover after period $\gamma_c^s$ which is multiplied by the rate of children that do recover from the disease $\rho_c$; 2) in each point, a portion of the asymptomatic infected children recover after period $\gamma_c^a$;

$$\frac{dR_c(t)}{dt} = \gamma_c^s \rho_c I_c^s(t) + \gamma_c^a I_c^a(t). \tag{S11}$$

In Eq. (S12), $\frac{dR_a(t)}{dt}$ is the dynamic amount of recovered adult individuals over time. It is affected by the following two terms: 1) in each point, a portion of the symptomatic infected adults recover after period $\gamma_a^s$ which is multiplied by the rate of adults that do recover from the disease $\rho_a$; 2) in each point, a portion of the asymptomatic infected adults recover after period $\gamma_a^a$.

$$\frac{dR_a(t)}{dt} = \gamma_a^s \rho_a I_a^s(t) + \gamma_a^a I_a^a(t). \tag{S12}$$

In Eq. (S13), $\frac{dD_c(t)}{dt}$ is the dynamic amount of dead children over time. It is affected by the portion of the symptomatic infected children that do not recover after period $\gamma_c^s$ which is multiplied by the rate of children that do not recover from the disease $1 - \rho_c$.

$$\frac{dD_c(t)}{dt} = \gamma_c^s (1 - \rho_c) I_c^s(t). \tag{S13}$$

In Eq. (S14), $\frac{dD_a(t)}{dt}$ is the dynamic amount of dead adult individuals over time. It is affected by a portion of the symptomatic infected adults that do not recover after period $\gamma_a^s$ which is multiplied by the rate of adults that do not recover from the disease $1 - \rho_a$.

$$\frac{dD_a(t)}{dt} = \gamma_a^s(1 - \rho_a)I_a^s(t). \tag{S14}$$
